# UV LED Wastewater Disinfection: The Future is Upon Us Full-scale installation and auditing of 280nm UV LED disinfection reactor

**DOI:** 10.1101/2024.06.12.24308830

**Authors:** Sean A MacIsaac, Bailey Reid, Carolina Ontiveros, Karl G Linden, Amina K Stoddart, Graham A Gagnon

## Abstract

The world’s first full-scale, 280 nm UV LED reactor for wastewater disinfection was tested at flows of 545 and 817 m^3^ day^-1^. The system achieved a >3 average log reduction of total coliform at 545 m^3^ day^-1^ and the 817 m^3^ day^-1^ flow rate achieved over a >2.5 average log reduction for all operational conditions. The delivered fluence of the full-scale system ranged from 28-148 mJ cm^-2^ and aligns with a UV auditing study that was conducted prior to the installation of the wastewater reactor. These results benchmark the performance that can be achieved by UV LED disinfection and further connect bench-scale disinfection results with full-scale performance. The approach established in this manuscript provides a novel tool for utilities when considering emerging UV disinfection technologies. In summary, this study establishes that UV LEDs are an effective wastewater disinfectant at-scale and are comparable to conventional low-pressure UV systems. This is the first instance where the efficacy of UV LEDs for municipal wastewater disinfection has been demonstrated using a large-scale installation at a functioning wastewater facility.

**Graphical Abstract:** 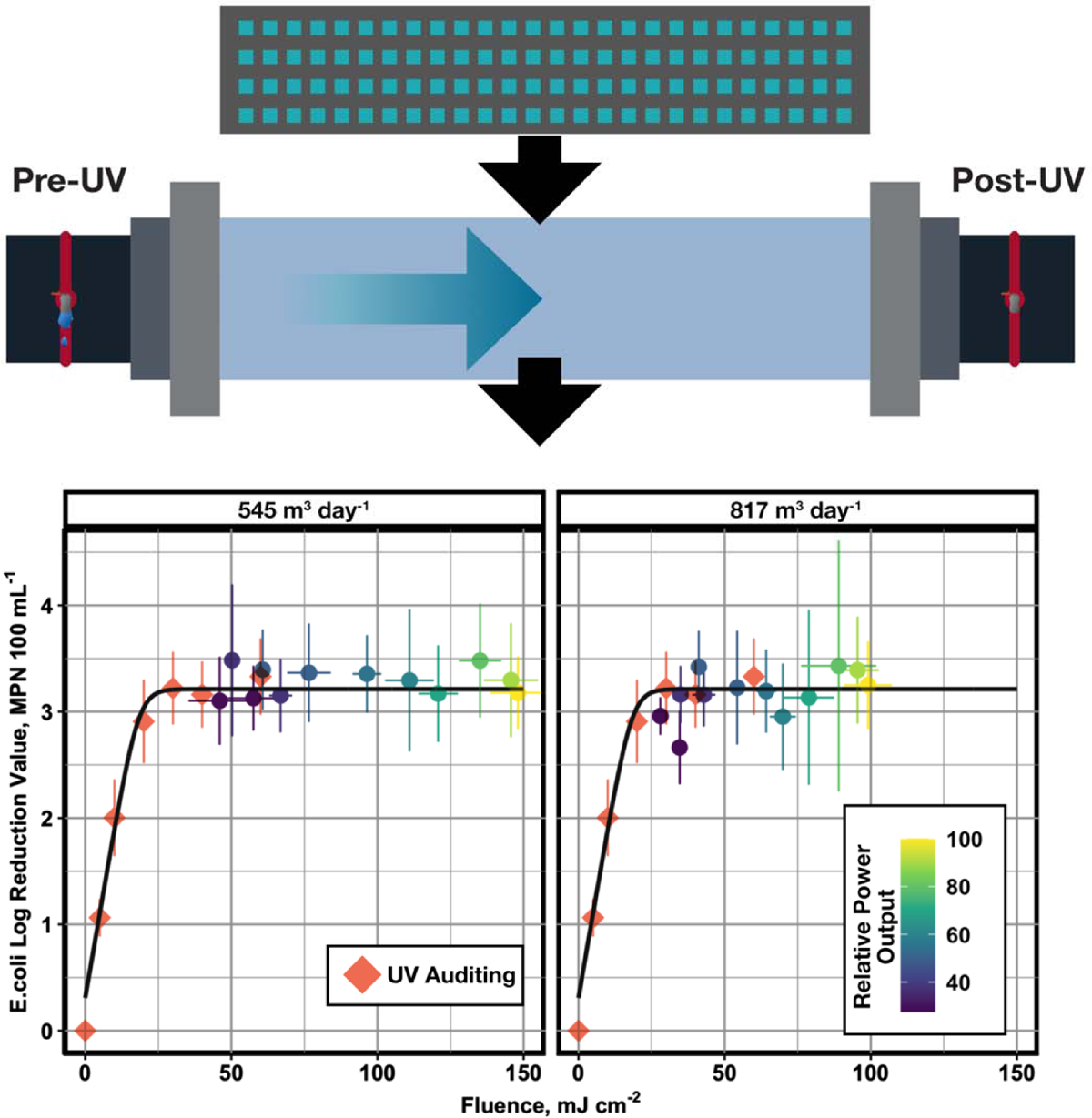

## Introduction

UV LEDs have been maturing as technology rapidly over the last ten years and have found numerous niche applications to date (Beck et al., 2017; Linden et al., 2019; Oguma, 2023). Many of these uses are for small-scale or point-of-use disinfection of pathogens (Beck et al., 2017; Jarvis et al., 2019; Oguma, 2023). Most of the research on UV LED disinfection has focused on drinking water applications, but UV disinfection is also commonly used in wastewater facilities. The overall market for industrial wastewater treatment is estimated to grow to a market cap of $16.5 billion by 2026 and the average global cost of wastewater infractructure is estimated to be upwards of $100 billion dollars (Dutta et al., 2021; Unesco, 2017). Given the sizeable market for treatment technologies, the efficiency of rapidly emerging UV LEDs for disinfection of wastewater matrices should be assessed. Recent work from MacIsaac, Rauch et al. demonstrated that not only are UV LED disinfection processes comparable to conventional low-pressure (LP) disinfection, but they can surpass them in certain circumstances in terms of disinfection efficacy at equivalent fluences (MacIsaac et al., 2023).

Conventional mercury-based systems pose a challenge in the coming years, where the intent to prohibit the handling of mercury is expected to come into effect between 2024 and 2028 (Government of Canada, 2023). Additionally, the future market for sourcing mercury is uncertain as mercury mining is set to be outlawed globally in 2032 (Bank, 2020; UN Environment Programme, 2023). UV LEDs present an alternative for these applications and have their own set of benefits. The modularity and compactness of UV LEDs enable the tailoring of reactors to footprint or geometry constraints (Sandhu, 2007). Diverse configurations of UV LEDs can impact disinfection efficacy, emphasizing the importance of optimizing reactor hydraulics and UV LED arrangements (Liu et al., 2023; Mohaghegh Montazeri and Taghipour, 2023). Additionally, the choice of wavelength is a pivotal factor in UV LED-based disinfection. While conventional UV mercury lamps are limited to 254 nm, recent studies have shed light on the distinct disinfection mechanisms of alternative wavelengths, such as 265 nm and 280 nm, against microorganisms (Martín-Sómer et al., 2023; Pousty et al., 2021; Song et al., 2016). These findings suggest that UV LEDs have unique applications that were not previously considered, but more research is needed to investigate this technology for full-scale applications.

This study investigates the first-ever full-scale UV LED reactor for wastewater treatment. The conditions observed in this research mark a significant jump from the next largest reactor described in peer-reviewed literature, which treated water at 185 GPM at a UVT_275_ between 90% and 97% (Jarvis et al., 2019). The reactor studied in this work had flows of 545 and 817 m^3^ day^-1^ (100 and 150 GPM) with a UVT_254_ of 50 - 60%. Here, we present new findings from a study that spans the installation, auditing, and full-scale testing of the world’s largest-capacity 280 nm UV LED wastewater reactor. This study addresses the paucity of information in the scientific literature concerning scaling UV LED technology and its suitability for challenging water matrices such as the comparatively lower UV transmittance (UVT) found in wastewater.

## Results and Discussion

### Water Quality Characterization

**Table 1** outlines the water quality parameters during the three sampling periods covered in this study. The pre-installation period ranged from June to October 2023, the dual panel testing from January to March 2024, and the single panel testing from March to April 2024. UVT measurements at 254 and 280 nm were collected as grab samples for every sampling event. Online UVT_254_ data was also gathered from the EPWWTF online sensor and is summarized in **Table 1**. Full-scale sensor readings were averaged on a per-day basis from continuous data spanning the course of each experimental period.

**Table 1.** Average water quality parameters during the test sampling periods.

| Parameters | Pre-Installation,<br>Conventional LP UV<br>Testing<br>(Summer 2023) | Dual UV LED<br>Panel Testing<br>(Winter 2024) | Single UV LED<br>Panel Testing<br>(Winter 2024) |
| --- | --- | --- | --- |
| UVT<sub>254</sub> Online<br>Sensor (%)* | 54.6 (SD= 5.5, n= 127) | 57.4 (SD= 6.7, n= 37) | 66.2 (SD= 3, n= 23) |
| UVT<sub>254</sub> grab (%) | 66.7 (SD= 4, n= 18) | 68.4 (SD= 4.3, n= 10) | 72.9 (SD= 4, n= 7) |
| UVT<sub>280</sub> grab (%) | 59.9 (SD= 4.3, n= 18) | 62.3 (SD= 4.8, n= 10) | 68.7 (SD= 3.6, n=7) |
| TSS grab (mg L<sup>-1</sup>) | 9.3 (SD= 9.5, n= 11) | 12.7 (SD= 13.1, n= 10) | 11.6 (SD= 3.7, n= 6) |
| Flow (m<sup>3</sup> day<sup>-1</sup>)* | 16,059 (SD= 8,259, n= 127) | 17,287 ( SD= 11,830, n=37 ) | 18,660 (SD= 8,461, n= 23) |
| UV Dose (mJ cm<sup>-2</sup>)* | 53.6 (SD= 9.4, n= 127) | N/A | N/A |
| Power (KW)* | 32.6 (SD= 11.9, n= 127 ) | N/A | N/A |

\*Daily averages from WW facility SCADA system

The analysis of wastewater quality data indicates significant differences in the daily average %UVT_254_ monitored online across the three testing periods (*p-value < 0.001*). Interestingly, the pre-installation and post-installation dual panel testing periods showed only marginal significance (*p-value = 0.061*). During the post-installation single panel period, the average percentage of UVT_254_ was higher at 66.2%, compared to 57.4% for the dual panel period and 54.6% for the pre-installation period. This trend was consistent when examining grab samples for both UVT_254_ and UVT_280_. Similarly, the daily average flow rate followed a similar pattern, showing overall statistical significance (*p-value = 0.02*). However, the pre-installation and post-installation dual panel testing periods did not significantly differ (*p-value = 0.62*). In contrast, TSS from grab samples did not demonstrate statistical differences across the three testing periods. Further details regarding observed water quality can be found in **SI Figure 5**.

### Pre-installation UV Auditing

The UV auditing approach (MacIsaac et al., 2023; Rauch et al., 2022) was used to estimate the full-scale LPUV system’s delivered fluence. **Figure 1** shows the pre-installation UV Auditing results for the EPWWTF wastewater during Summer 2023. The boxplots on the plot represent the bench-scale disinfection data for 254 and 280 nm UV exposures, and the dashed line is the mean log reduction for the full-scale LPUV system. The point where the boxplots intersect the full-scale performance provides an indication of the fluence that is achieved by the full-scale system. This dataset suggests that the EPWWTF achieves a ≈2.5-log reduction at a fluence of approximately 20 mJ cm^-2^. This means that the full-scale facility could be acheviing equivalent log reduction performance at half of the operational fluence that was measured during the pre-installation period described in **Table 1**.

**Figure 1.**
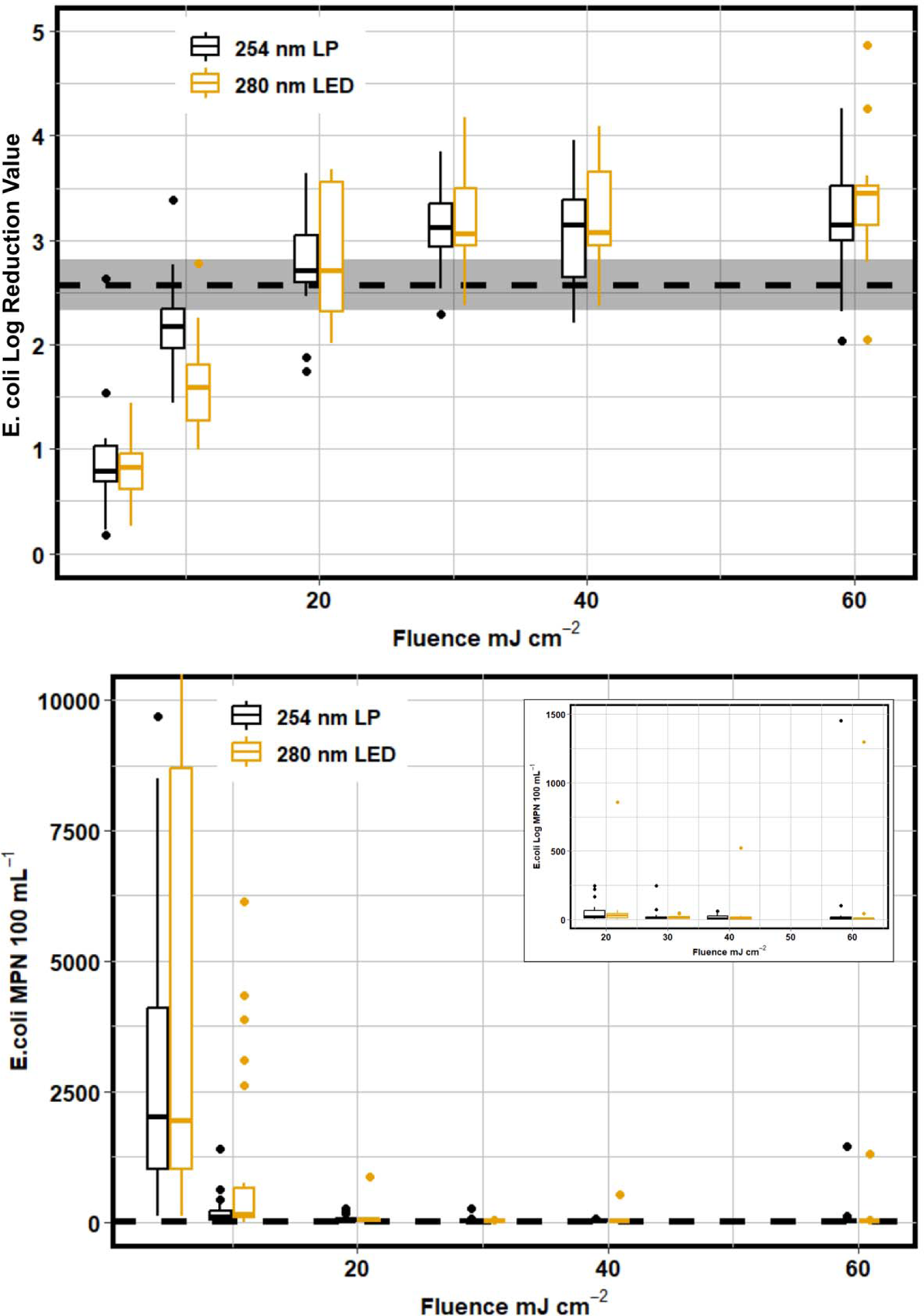
Pre-installation UV auditing data for the EPWWTF comparing bench-scale LP and LED inactivation to full-scale conventional disinfection (n=19). The dashed line with grey shaded region represents the full-scale conventional system performance and 95% confidence interval about the mean log reduction value.

As shown in Figure 1, UV LEDs at 280 nm were comparable to conventional disinfection at 254 nm across all tested fluences, which differs from previously published data that showed that LEDs can outperform conventional disinfection (MacIsaac et al., 2023). This result highlights the importance of characterizing the water matrix targeted for disinfection when considering LED technologies. Both 254 nm and 280 nm collimated beam data indicated that the ceiling of disinfection is approximately 3.5 log, which suggests that this specific wastewater matrix is matrix limited beyond a fluence of 30 mJ cm^-2^. The tailing portion of the disinfection curve begins at a fluence of 20 mJ cm^-2^ and shows little difference in log reduction value between 30 and 60 mJ cm^-2^. This means that the full-scale facility is relatively optimized given their specific wastewater quality conditions, and it is unlikely that improvements above a 3-log reduction for the conventional full-scale system could be achieved. However, this data shows that LEDs can compete with conventional 254 nm disinfection across various water qualities.

### UV LED Reactor Post-Installation Results – Dual and Single Panel Results

Based on this specific reactor design, the post-installation data shows that 280 nm disinfection can be scaled appropriately to a full-scale operation. Figure 2 depicts the full-scale performance of UV LEDs compared to the average log reduction value for the full-scale conventional system. Every operational condition tested for both 545 and 817 m^3^ day^-1^ flow rates achieved a log reduction between 2.8 and 3.8. This result indicated that the full-scale UV LED reactor overperformed what was anticipated prior to installation.

**Figure 2.**
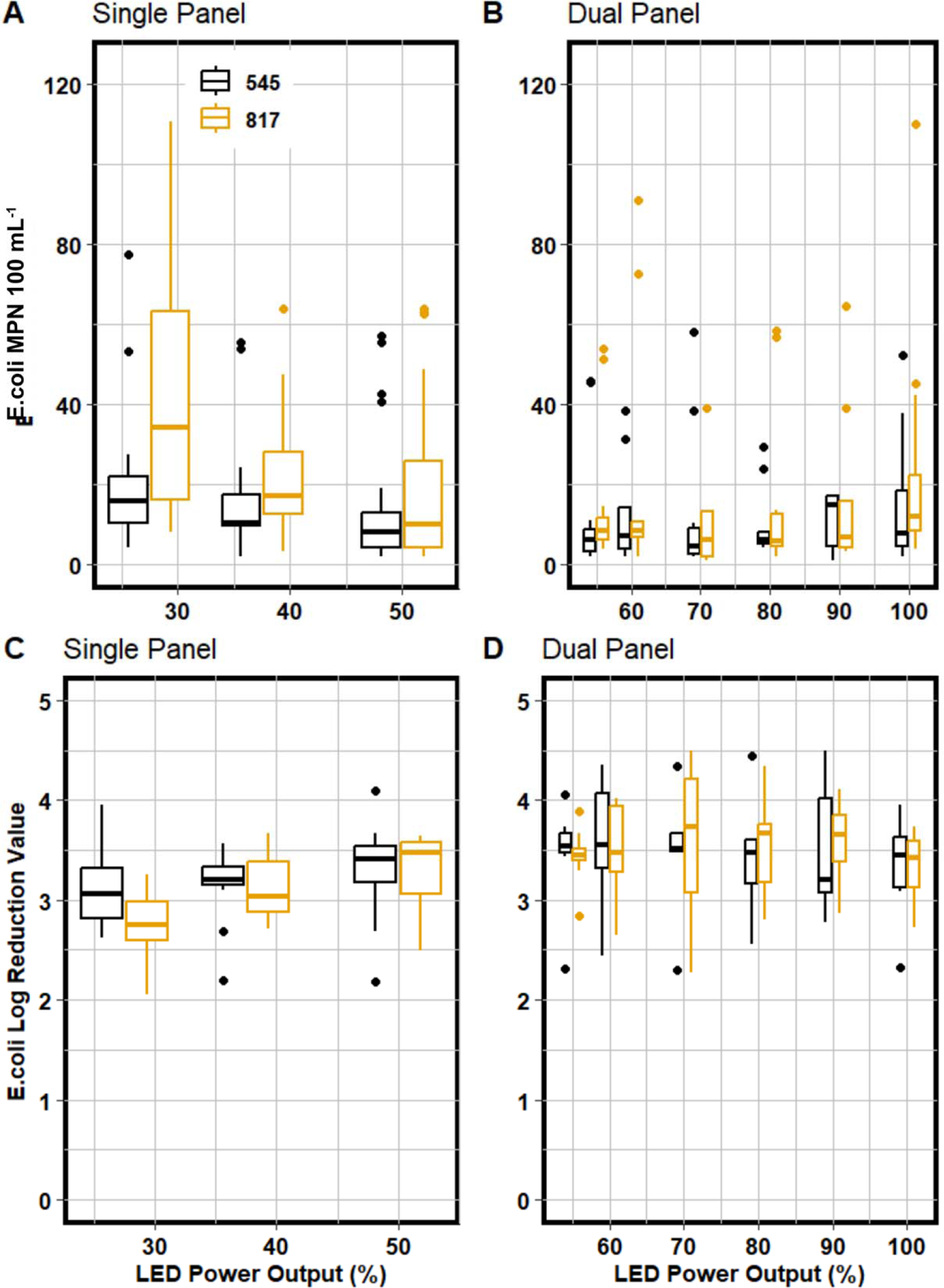
Full-scale 280 nm UV LED performance data for 545 and 817 m^3^ day^-1^ for single and dual panel UV LED operation. The top row shows the MPN 100 mL^-1^counts vs the bottom row which shows the relative log reduction values for each of the operational conditions.

During the post-installation single panel testing period, the UV LED reactor had an average log reduction value (LRV) of 3.2 when run at 545 m^3^ d^-1^ and 817 m^3^ d^-1^ when operated at 100% LED power outputs. There was only a marginal difference in LRVs between the two flows at the 27.5% power output (*p-value = 0.031*); in the rest of the UV LED Power Outputs (%) tested, there was no difference between the two flows. Furthermore, at 545 m^3^ d^-1^, there was no difference between all of the LED Power Outputs (%) (*p-value = 0.48*), whereas, at 817 m^3^ d^-1^, only 27.5 % resulted different from the rest of the levels with a mean LRV of 2.81 ± 0.3 (*p-value = 0.009*). In contrast, the other UV LED Power Outputs achieved a mean LRV of 3.2 ± 0.09 (*p-value = 1*). When considering all factors together, there was no significant difference (p-value=0.079) in LRV between both flows and the various UV LED Power Outputs (%) tested. Further details can be found in **SI** Figures 6 **and 7**.

Based on the pre-installation UV auditing data gathered during the Summer of 2023, the UV LED full-scale reactor consistently delivered a UV fluence of at least 30 mJ cm^-2^. The delivered fluence was further investigated by taking advantage of the online UV intensity data and the UVT_280_ collected via grab samples during the collection period.

A detailed approach for estimating the full-scale reactor’s fluence relied on the fluence calculations described by Bolton and Linden (2003). This approach relies on utilizing the online UV intensity sensors that are equipped on the bottom of the UV LED reactor. Utilizing this value inherently incorporates changing UVT within the wastewater matrix, as less light will reach the sensor during periods of low UVT. This approach assumes a simple residence time of influent water by calculating time using the online flow data and the dimensions of the cylindrical reactor chamber. This approach is summarized in **Equation 1** and **Equation 2**.

Equation 1 Average irradiance for the full-scale UV LED reactor

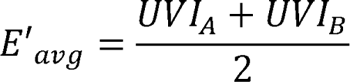

E^’^_avg_ = Average UV irradiance for each of the online sensors

UVI_A_ = Online UV irradiance for panel A

UVI_B_ = Online UV irradiance for panel B

Equation 2 Mean fluence calculation for estimating the delivered fluence via the full-scale UV LED reactor

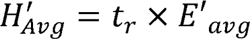

H^’^_avg_ = Fluence estimation using manufacturer method

t_r_ = Residence time of influent wastewater

Calculating a delivered fluence is essential when considering the energy needs and efficiencies of UV LEDs compared to conventional UV treatment. Fluence was estimated using the volumetric mean flow and volumetric mean fluence rate using **Equation 2**. The residence time was determined using the online flow data and volume of the reactor chamber, whereas the average irradiance was determined using paired UV intensity data from each of the two sensors equipped with the reactor. The fluence was then calculated for each sampling event and operating condition captured in the ten-week period. A further assessment of the fluence approach was done by pairing each calculated fluence with the corresponding log reduction values for the single and dual-panel UV LED conditions. This data was then directly compared to the UV auditing data to understand how the fluences at bench and full-scale compared to log reduction values.

The unique operational conditions were binned together so that 95% confidence intervals for fluence and log reduction could be calculated. The resulting plot is shown in Figure 3. The colour ramp for this plot is normalized to the total nominal power setting (i.e. 100% represents both LED panels set to 100% output, and 27.5% represents a single UV LED panel set to 55%). The top row of this Figure displays the log reduction values for both pre-installation UV auditing and full-scale sampling. The 95% confidence intervals show a minimum log reduction of 2.66, achieved when a single UV LED panel is set to the minimum (55%) power output setting. This corresponds to a delivered fluence of 28.0 ± 2.78 mJ cm^-2^ for single panel output at a flow rate of 817 m^3^ day^-1^. The minimum delivered fluence for the 545 m^3^ day^-1^ is 46.0 ± 10.7 mJ cm^-2^. These values are in alignment with the pre-installation UV auditing data that suggests that the matrix limitation of the EPWWTF is reached at a fluence of approximately 30 mJ cm^-2^.

**Figure 3.**
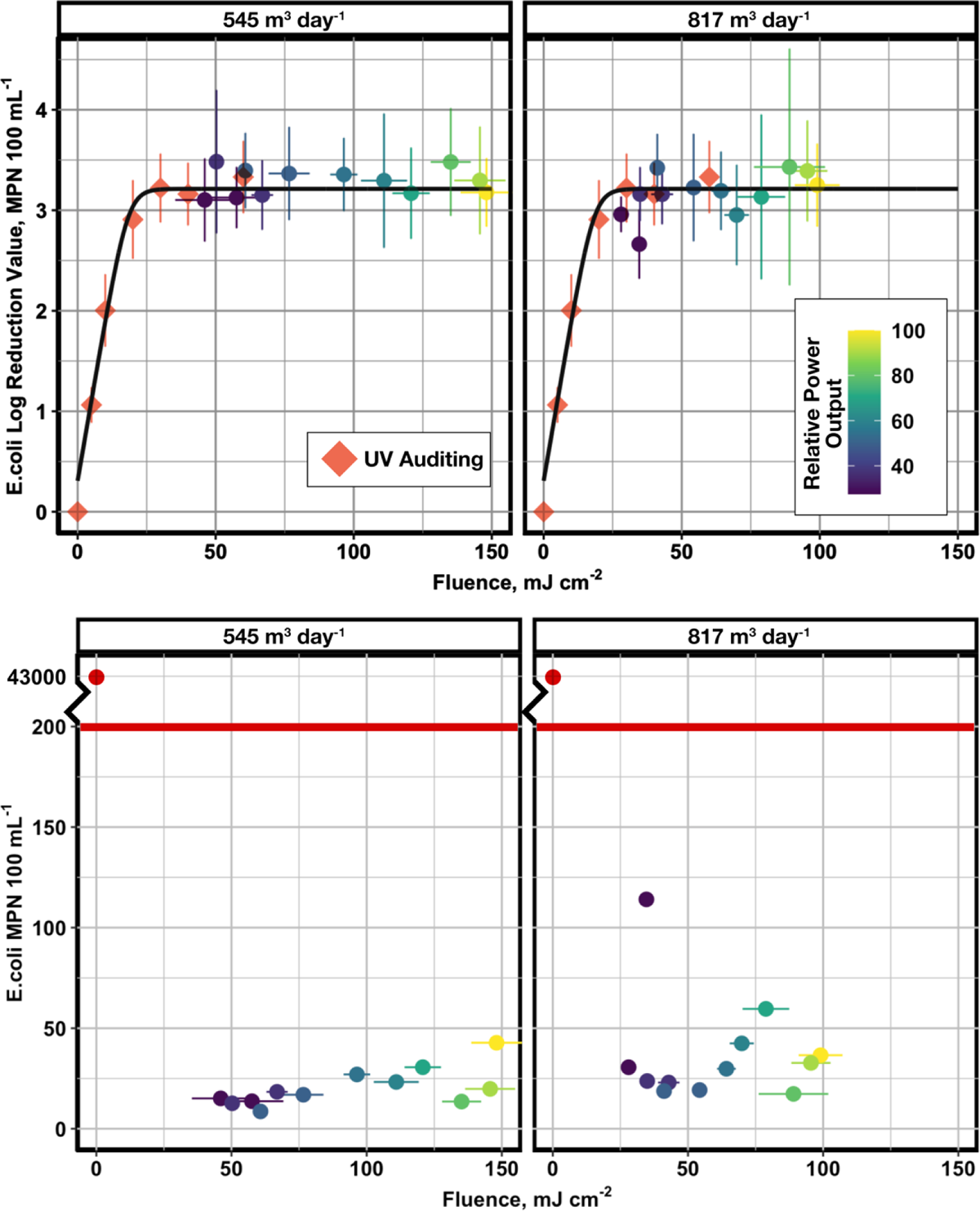
Fluence estimation for each normalized power setpoint for single and dual UV LED panel disinfection and faceted for each operational flow rate. Error bars represent the 95% confidence intervals. The colour ramp indicates the UV LED system’s normalized, relative power output.

The bottom row of Figure 3 shows the MPN 100 mL^-1^ values for full-scale sampling. The red line indicates the regulatory threshold for the EPWWTF and the axis has been truncated to properly show the differences in treated samples when compared to the untreated influent wastewater. This shows that the UV LED reactor performed below the regulatory threshold for all conditions.

Geeraerd’s model was applied to the pre-installation UV auditing dataset to determine the inactivation kinetics specific to the EPWWTF. Geeraerd’s model was chosen as it has extensive use in other UV LED studies and is able to capture tailing effects in wastewater matrices (Geeraerd et al., 2000; Rattanakul and Oguma, 2018; Rauch et al., 2022). The k-value for the EPWWTF was determined to be 0.160 cm^2^ mJ^-1^ and aligned with the full-scale datasets. This result indicates that the approach for calculating the delivered fluence for the full-scale system is a valid estimate. This outcome establishes that the UV auditing approach can be paired with full-scale UV LED piloting to enhance the operating conditions for full-scale systems. This is a powerful tool for optimizing these systems and fine-tuning the brightness of LED panels.

## Conclusion

In summary, this study demonstrated a world’s first in utilizing UV LEDs for full-scale wastewater disinfection across 24 different operational conditions. These operating conditions showed that UV LEDs are capable of being used for the disinfection of wastewater. A 3-log reduction was observed for all operating conditions at an average UVT_254_ ranging from 54.6% to 66.2%, and an average UVT_280_ ranging from 59.9% to 68.7%. The assessment of fluence suggests that the reactor achieved a minimum fluence of approximately 28 mJ cm^-2^ at a setpoint of 55% for a single panel and a flow rate of 817 m^3^ day^-1^. This result aligns with the pre-installation, UV auditing data, demonstrating that UV disinfection for this specific wastewater matrix approaches a tailing, matrix-limited region at fluences of at least 30 mJ cm^-2^.

In this work, the 280nm UV LED reactor inactivated significantly more *E. coli* and total coliforms than anticipated which prevented a more precise assessment of the reactor performance limitations. All full-scale UV exposure were near or in the tailing region of disinfection, but aligned with bench-scale auditing. Future work is planned to evaluate UV LED performance at higher flow rates and lower power outputs to achieve fluences within the log linear portion fo the disinfection curve. Overall, this work demonstrates that UV LED technologies can be applied at a municipal-scale use and are not only for small-scale applications in drinking water treatment.

## Materials and Methods

The work described in this study covers two main sampling periods, referred to as pre-installation, compiled during the Summer of 2023 and post-installation for single and dual panels, which was collected during the Winter of 2024.

### Pre-Installation Sample Collection

Eastern Passage Wastewater Treatment Facility (EPWWTF) is a conventional activated sludge wastewater treatment facility with average daily flows of approximately 5,448 m^3^d^-1^. UV auditing, as described by Rauch et al. (2022) and MacIsaac et al. (2023), using EPWWTF wastewater was conducted twice weekly over the course of four months during summer 2023 (June – October), totalling 19 sampling events during this period. Sampling from two points in the wastewater treatment process was included in the UV auditing. The first sampling point was prior to UV disinfection and after secondary clarification (Pre-UV). The second sampling point was after UV disinfection from the open-channel, full-scale, conventional disinfection system (Post-UV). Samples from both locations were collected in 1 L bottles, placed in coolers, and transported to Dalhousie University to be analyzed. The delivered fluence from the full-scale conventional UV system was estimated using the UV Auditing approach.

A low-pressure UV (LPUV) collimated beam unit (Calgon Carbon Corporation, PA, USA) and a UV LED collimated beam ( PearlLab Beam AquiSense, Erlanger, KY, USA) were used for conventional and LED bench-scale exposures, respectively. UV fluences of 5, 10, 20, 30, 40, and 60 mJ cm^-2^ were used for both 254 nm (conventional) and 280 nm (UV LED) exposures and calculated following the approach described by Bolton and Linden (2003).

Pre-UV samples were analyzed for UVT, total suspended solids (TSS), *E. coli* and total coliforms, whereas post-UV samples were only analyzed for *E. coli* and total coliforms. TSS was determined according to standard method 2540 D (American Public Health Association et al., 2018). UVT% at 254 and 280 nm were measured on a spectrophotometer (DR 5000 Spectrophotometer, HACH Company, Loveland, CO, USA). *E. coli* and total coliforms were enumerated using a commercial multiwell enzyme substrate test (Rodger and Bridgewater, 2017). *E. coli* and total coliform most probable number (MPNs) were used to develop fluence response curves during the auditing period. This auditing was conducted before installing the UV LED full-scale reactor.

### UV LED Full-Scale Reactor Installation

The commissioning and installation of the full-scale UV LED reactor (Tera 280 nm UV LED reactor, AquiSense, Erlanger, KY, USA) immediately followed the pre-installation phase. The reactor was fully integrated into the Supervisory Control and Data Acquisition (SCADA) system at the EPWWTF. This setup enabled dynamic control over the flow rate and the power output of the UV LED banks and logged lamp performance continuously during the study period. The specific reactor arrangement used in this study allowed for power outputs ranging from 55% to 100% for each of the two 280 nm UV LED panels. Figure 4 shows the reactor installed at the EPWWF. The pump used for this work delivered flow rates ranging from 545 and 817 m^3^ day^-1^.

**Figure 4.**
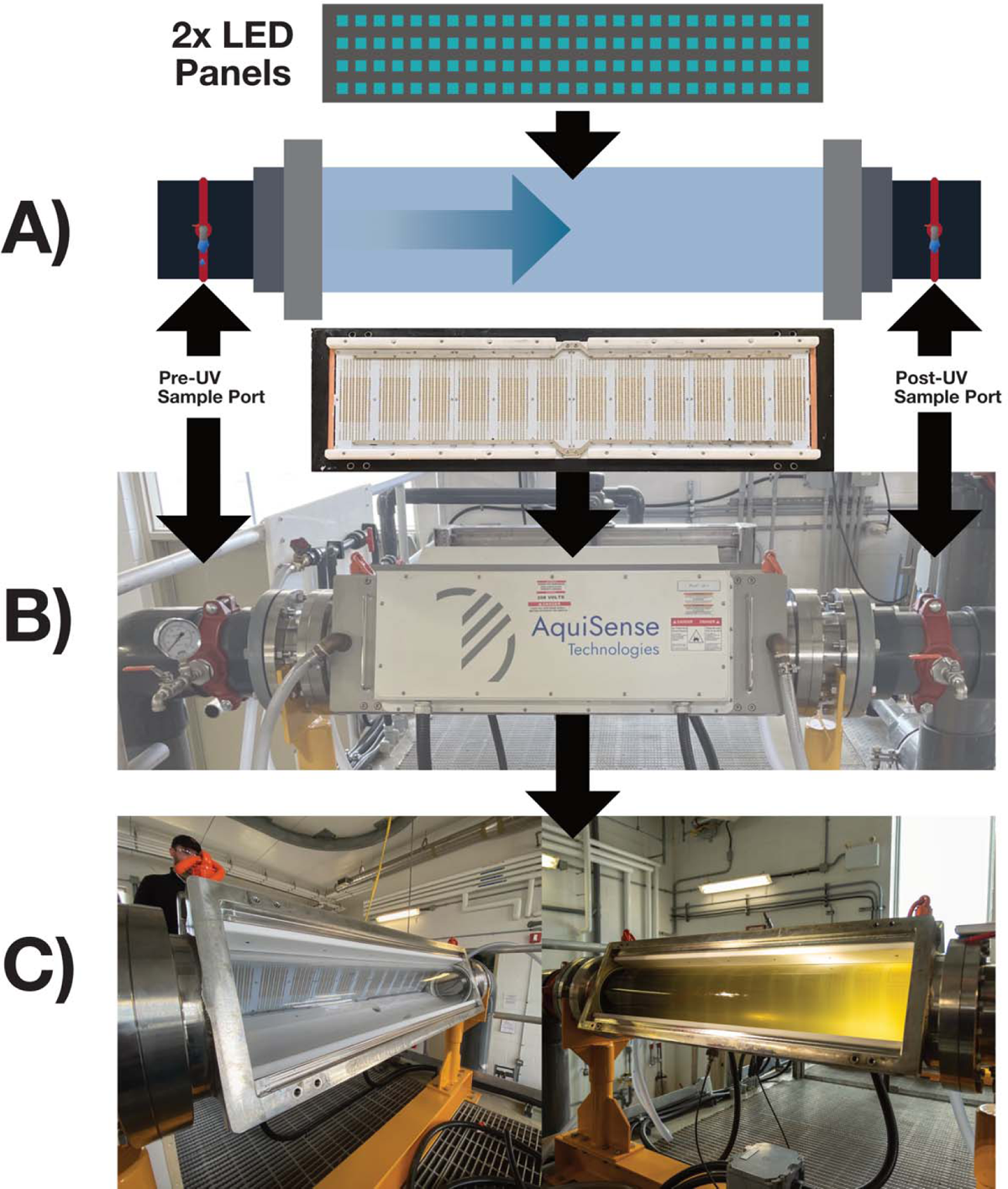
Schematic of the 280 nm UV LED reactor. A) overview of sampling ports at the influent and effluent of reactor; B) photograph of the installed reactor at the EPWWTF; C) View of the internal reactor chamber before and after opening the influent wastewater flow valve

### UV LED Full-Scale Sampling

Full-scale reactor performance sampling was conducted at the EPWWTF twice weekly for a 10-week period during Winter 2024 (January-April), totalling ten sampling events. Samples were collected from three points in the wastewater treatment process: pre-UV disinfection, post conventional UV disinfection, and post UV LED disinfection. Sampling from the conventional system followed the method developed for the pre-installation phase. The sampling immediately after the UV LED was collected for various operational conditions for the UV LED panels. The first five weeks of sampling consisted of setting both LED panels to equivalent relative power outputs of 55, 60, 70, 80, 90 and 100% and two set flow rates of 545 and 817 m^3^ day^-1^. 55% relative power output was the lowest possible setting for this specific reactor. The second five-week period consisted of independent control of the UV LED panels to understand how single-panel output impacted disinfection. Each panel was independently set to 0, 55, 75, and 100%, respectively, for sampling.

Colilert tray counts were used for all biological quantification during the sampling period according to standard methods (Rodger and Bridgewater, 2017). This approach was previously described by Rauch et al. (2022) when assessing full-scale wastewater facilities. Pre-UV UVT _254_, TSS, and flow rate were also monitored as part of regular grab sampling and the online SCADA system at the treatment facility.

## Data Availability

All data produced in the present study are available upon reasonable request to the authors

## Acknowledgments

The authors would like to acknowledge the support from the NSERC Alliance program (Grant Number: (ALLRP 568507-21). The authors would also like to acknowledge the support from the Water Research Foundation as this work is part of WRF Project #5173 which investigates the feasibility of UV LED technologies for water treatment.

## Data Availablity

Data collected in this study is available upon request from the authors.

## Notes

### Competing Interest Statement

The authors have declared no competing interest.

### Funding Statement

This study was funded by Water Research Foundation Project 5173

